# Diagnostic performances and thresholds: the key to harmonization in serological SARS-CoV-2 assays?

**DOI:** 10.1101/2020.05.22.20106328

**Authors:** Mario Plebani, Andrea Padoan, Davide Negrini, Benedetta Carpinteri, Laura Sciacovelli

## Abstract

**Background:** The evaluation of severe acute respiratory syndrome coronavirus 2 (SARS-CoV-2) specific antibody (Ab) assay performances is of the utmost importance in establishing and monitoring virus spread in the community. In this study focusing on IgG antibodies, we compare reliability of three chemiluminescent (CLIA) and two enzyme linked immunosorbent (ELISA) assays.

**Methods:** Sera from a total of 271 subjects, including 64 reverse transcription-polymerase chain reaction (RT-PCR) confirmed SARS-CoV-2 patients were tested for specific Ab using Maglumi (Snibe), Liaison (Diasorin), iFlash (Yhlo), Euroimmun (Medizinische Labordiagnostika AG) and Wantai (Wantai Biological Pharmacy) assays. Diagnostic sensitivity and specificity, positive and negative likelihood ratios were evaluated using manufacturers’ and optimized thresholds.

**Results:** Optimized thresholds (Maglumi 2 kAU/L, Liaison 6.2 kAU/L and iFlash 15.0 kAU/L) allowed us to achieve a negative likelihood ratio and an accuracy of: 0.06 and 93.5% for Maglumi; 0.03 and 93.1% for Liaison; 0.03 and 91% for iFlash. Diagnostic sensitivities and specificities were above 93.8% and 85.9%, respectively for all CLIA assays. Overall agreement was 90.3% (Cohen’s kappa = 0.805 and SE = 0.041) for CLIA, and 98.4% (Cohen’s kappa = 0.962 and SE = 0.126) for ELISA.

**Conclusions:** The results obtained indicate that, for CLIA assays, it might be possible to define thresholds that improve the negative likelihood ratio. Thus, a negative test result enables the identification of subjects at risk of being infected, who should then be closely monitored over time with a view to preventing further viral spread. Redefined thresholds, in addition, improved the overall inter-assay agreement, paving the way to a better harmonization of serologic tests.

## INTRODUCTION

The spread of coronavirus disease 2019 (COVID-19), caused by the severe acute respiratory syndrome coronavirus 2 (SARS-CoV-2), has become a pandemic, with sustained human-to-human transmission. Since the initial identification of COVID-19 in December 2019, there has been an exponential rise in the number of cases worldwide. The reasons for the rapid spread include the high transmissibility of the virus, especially among asymptomatic or minimally symptomatic carriers, as well as the apparent absence of any cross-protective immunity from related coronavirus infections, and the tardy public health response measures (1-2).

An accurate diagnosis of SARS-CoV-2 infection is required for prompt and effective patient care. In particular, the rapid identification of cases among hospitalized patients remains a high priority in assuring prompt, and effective, treatments, allocating personal protective equipment (PPE), and in preventing nosocomial spread with subsequent community transmission. In addition, accurate diagnosis is of paramount importance in controlling the outbreak, establishing protective measures, monitoring therapy and conducting epidemiological surveillance (3). The detection of the viral genome in respiratory samples, particularly nasopharyngeal specimens for swab-based SARS-CoV-2 testing with RT-PCR, is currently considered the “gold standard” for confirming a clinically suspected diagnosis and identifying asymptomatic carriers (4). COVID-19 infection can also be detected indirectly by measuring the host immune response to SARS-CoV-2 infection. Virus-specific antibody (Ab) detection for COVID-19 should complement nucleic acid testing, particularly in the later stages of infection (i.e., when the virus has been eliminated) (5), in surveying for asymptomatic infection in close-contacts, in establishing and monitoring the extent of viral spread in the community, and in conducting epidemiological surveillance (4).

The several assays developed and currently available on the market, differ due to two major variables: a) the format used and, in particular, whether the test is a quantitative laboratory-based immunoassay ELISA, CLIA or a qualitative point-of-care test (POCT); and b) the SARS-CoV-2 antigen targeted, in particular if the antibodies are addressed against the spike surface protein (s) (namely subunit 1 and 2), and/or the spike receptor binding domain (RBD), and/or the nucleocapside protein (NC) (6). In order to appropriately use serological tests “for the right patient at the right time”, it is therefore important to validate serological methods that can be used in a specific patient as well as in large-scale studies, by comparing different available methods in order to identify the right cut-off as well clinical performance and diagnostic accuracy.

Aim of this study was to evaluate different chemiluminescent (CLIA) and enzyme-linked immunosorbent (ELISA) assays for SARS-CoV-2 antibodies in COVID-19 patients and healthcare operators, to identify appropriate cut-offs and evaluate diagnostic accuracy.

### Materials and Methods

#### Patients

A total of 271 subjects (87 healthcare workers, 64 SARS-CoV-2 patients, 19 autoimmune patients and 101 blood donors) underwent analysis with the different chemiluminescent (CLIA) and enzyme linked immunossays (ELISA) systems in order to verify immune response in relation to the different subject categories. With the exception of donors, all other subjects underwent at least one nasopharyngeal swab test, analyzed by RT-PCR. Of the 87 healthcare workers, 71 were considered negative (Neg-HW), since at least three sequential molecular results obtained between February 26th and April 10th, 2020 were negative, and the remaining 16 were considered positive with mild disease (Mild), and recovered at home, with supportive care and isolation. The 64 SARS-CoV-2 patients had moderate or severe disease and were monitored throughout hospitalization; 32 recovered with, and 32 without, the need of air ventilation support (Mod and Sev, respectively). The 19 autoimmune patients, who were SARS-CoV-2 negative and regularly referred to the clinic’s consultants for autoimmune disease (AI), were included in order to evaluate possible analytical interferences. The 101 donors were included because specimens had been collected from them and frozen at -80°C in 2015, before the emergence of SARS-Cov-2 (Pre-COV). Table 1 reports on the number and characteristics of subjects included in the study for gender, age and negativity or positivity to the virus SARS-CoV-2.

**Table 1.**
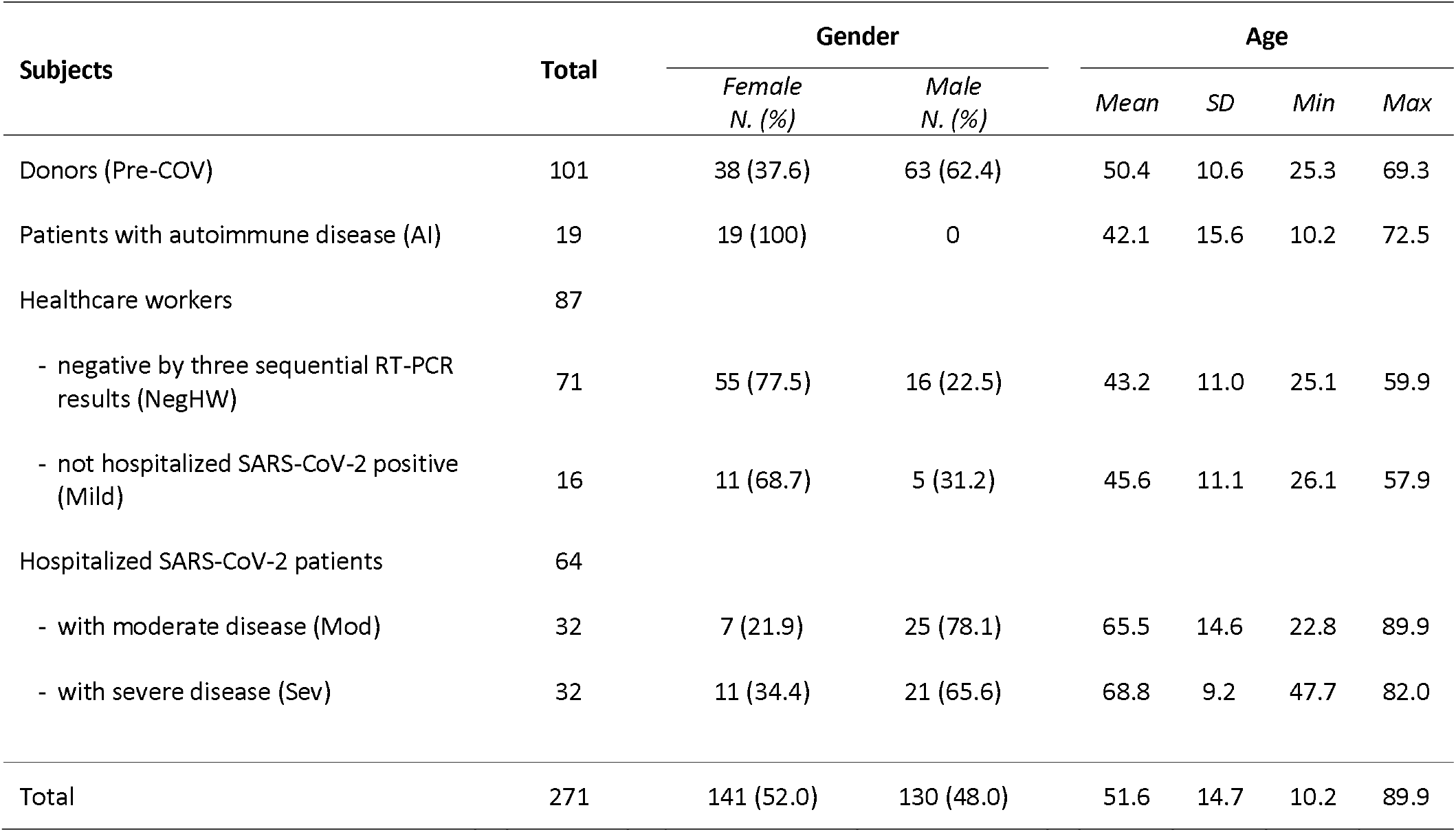
- Demographic characteristics of subjects included in the study.

The study protocol (number 23307) was approved by the Ethics Committee of the University-Hospital, Padova.

#### Analytical systems

The following five different analytical systems were evaluated.

- Maglumi™ 2000 Plus (New Industries Biomedical EngineeringCo., Ltd [Snibe], Shenzhen, China), a chemiluminescent analytical system (CLIA) for the detection of both IgM and IgG antibodies against SARS-Cov-2 S-antigen and N-protein. The analytical performances of this system are reported elsewhere (7). According to the manufacturer’s inserts (271 2019-nCoV IgM, V2.0, 2020-03 and 272 2019-nCoV IgG, V1.2, 2020-02), the clinical sensitivities of IgM and IgG were 78.65% and 91.21%, respectively, while the specificities of IgM and IgG were 97.50% and 97.3%, respectively.
- Liaison® SARS-CoV-2 S1/S2 IgG [DiasorinS.p.A, 13040 Saluggia (VC) – Italy], the fully automated indirect chemiluminescent immunoassay serology test for detecting IgG antibodies against S1/S2 antigens of SARS-CoV-2. According to the manufacturer’s inserts (Ref. 311450 EN-200/007-797, 01-2020-04), the IgG cut-off is: negative, <12.0kAU/L; equivocal, from 12 kAU/L to 15.0 kAU/L; positive, ≥15.0 kAU/L.
- iFlash-SARS-CoV-2 (Shenzhen Yhlo Biotech Co. Ltd.), a paramagnetic particle chemiluminescent immunoassay (CLIA) for the determination of IgM and IgG antibodies against SARS-CoV-2 nucleocapsid protein and spike protein. According to the manufacturer’s inserts (V1.0 English Fd. 2020-02-20), the IgM and IgG cut-off is 10.0 kAU/L.
- Euroimmun (MedizinischeLabordiagnostika AG – Lubeck, Germany) immunoassays (ELISA) for the detection of anti-SARS-CoV-2 IgA and IgG against SARS-CoV-2 spike protein. According to the manufacturer’s insert (Ref. EI_2606G_A_IT_C01), the IgG cut-off is: <0.8 ratio, negative; from 0.8 to 1.1 ratio, borderline; > 1.1 ratio, positive, as reported elsewhere (8).
- Wantai SARS-CoV-2 Ab Rapid Test (Beijing Wantai Biological Pharmacy Enterprice Co. LdtBioMedomicsInc,) an enzyme-linked immunosorbent assay (ELISA) for the detection of total antibodies (AbT) binding SARS-CoV-2 spike protein receptor domain (RBD) (6). The cut-offs are mathematically calculated in each run.

#### Repeatability and intermediate precision evaluation of CLIA assays

For Liaison and iFlash, precisions were estimated by using two or three human serum pools, respectively, of samples with different values. Estimations of precision were obtained by means of triplicate measurements of aliquots of the same pool, performed for a total of five consecutive days (Liaison) or three consecutive days (iFlash). Analysis of variance was used to estimate precision. Maglumi data on repeatability and intermediate precision are reported elsewhere (7).

#### Statistical analyses

Analyses were performed using Stata v13.1 (StataCorp, Lakeway Drive, TX) and R software v 4.0 (The R Foundation for Statistical Computing) and MedCalc Statistical Software version 19.2.1. Mean and standard deviations were used for descriptive statistics. Logarithmic transformation (log_10_) was applied to skewed data when necessary, before using parametric t-test to compare groups. Multiple comparisons were made using the calculation of Bonferroni adjusted (B-adj) p-values. Fisher’s exact test was employed to evaluate categorical data. The empirical method and Youden index were used to estimate the area under (AUC) the receiver operating characteristics curve (ROC) and best thresholds, respectively. Bland Altman analysis and Passing-Bablok regressions were used to assess the comparability of CLIA assays. Assessment of agreement was performed by concordance (in percentage) and by Cohen’s kappa. Thresholds for harmonizing assay results were determined by an in-house R script iterating the assessment of agreement for all the possible combinations of methods cut-offs, considering a minimum incremental delta of 0.2 kAU/L.

### Results

#### Patients’ characteristics

Table 1 reports the demographic characteristics of the study subjects/ healthcare workers, SARS-CoV-2 patients autoimmune patients and donors. The (total) overall mean age of subjects was 51.6 years with a standard deviation (±SD) of 14.7 (min, 10.2; max, 89.9); 141 (52%) were females. Age differed among: Pre-COV and Neg-HW (B-adj p = 0.0003) or Mod (B-adj p < 0.0001) or Sev (B-adj p < 0.0001); AI vs Mod (B-adj p < 0.0001) or Sev (B-adj p < 0.0001); Neg-HW vs Mod (B-adj p < 0.0001) or Sev (B-adj p < 0.0001); Mild vs Mod (B-adj p < 0.0001) or Sev (B-adj p < 0.0001). In the groups studied, there was no significant difference between the average age of females and that of males. Overall, the percentage of females and that of males differed significantly (p< 0.001), in particular for AI patients. For SARS-CoV-2 patients, the mean time interval from the onset of symptoms and serological determinations was 24 days (SD ±11; range 12 - 54 days).

#### Repeatability and intermediate precision evaluation of CLIA assays

The results of Liaison precisions calculated for IgG at two levels for repeatability and intermediate precision were: 3.5% and 4.5%, respectively at the level of 1.6 kAU/L, 4.0% and 7.9% at the level of 28.0 kAU/L. iFlash precisions were calculated for IgM and IgG at three different levels. For IgM, the results for repeatability and intermediate precision were: 8.7% and 9.4% at level of 2.1 kAU/L, 1.2% and 10.8% at level of 8.4 kAU/L, 5.1% and 11.4% at 75.3 kAU/L. For IgG, repeatability and intermediate precision results were: 3.7% and 3.7% at a level of 3.8 kAU/L, 2.1% and 2.3% at a level of 21.1 kAU/L, 1.3% and 3.6% at a level of 111.5 kAU/L. Only the iFlash insert reported data on precision (repeatability < 10% and intermediate precision < 15%).

#### Ab determined in different assays

IgG and IgM Ab results are reported in Figures 1 and 2, a log_10_ scale being used to enhance the visualization of data dispersion and box plots. Donors (only for CLIA assays) and negative autoimmune patients’ results were included to verify possible analytical interferences and differences with respect to healthcare workers who repeatedly tested negative to nasopharyngeal swab. Manufacturers’ cut-offs are reported in Table 2.

**Figure 1.**
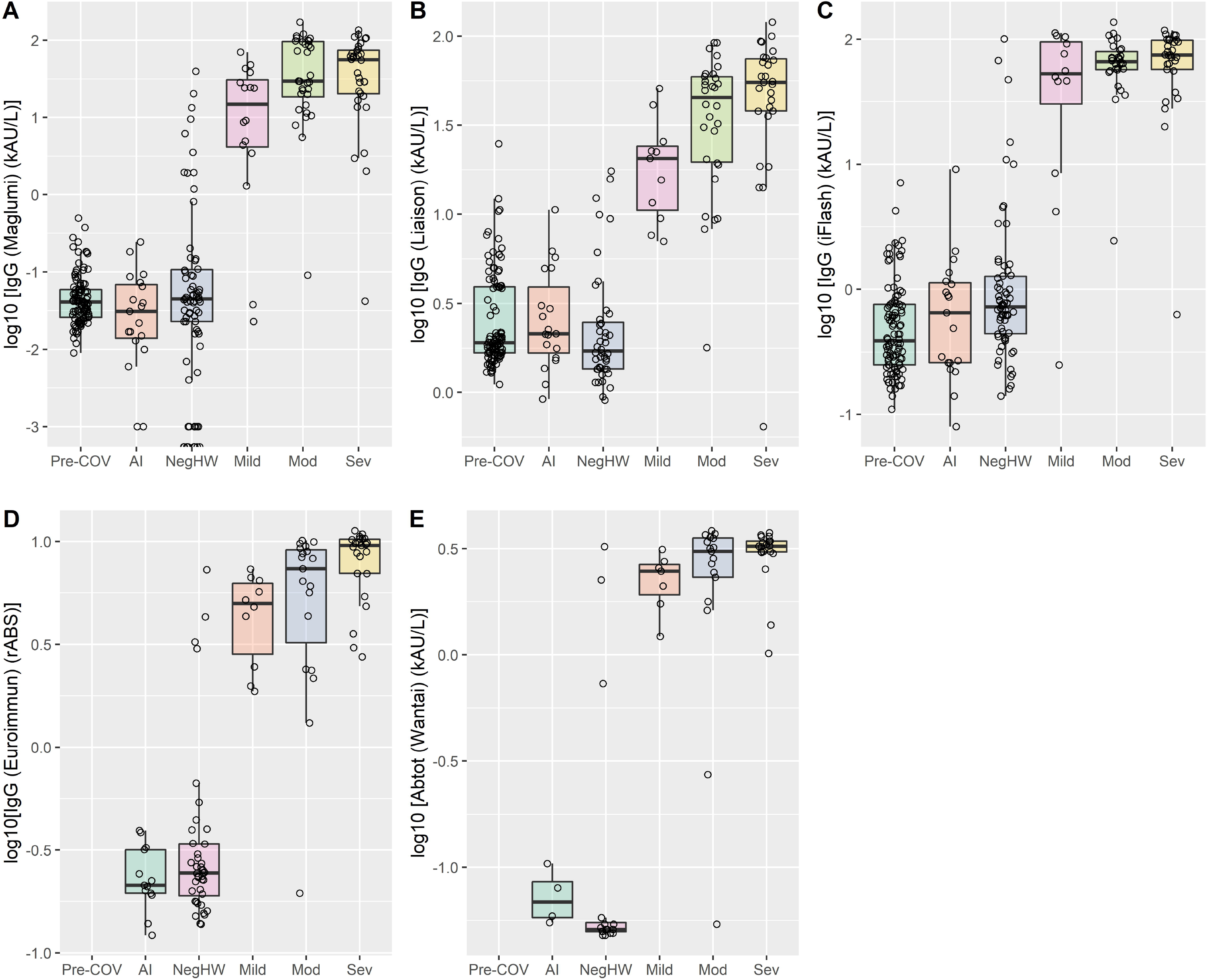
– IgG antibody levels for Maglumi (A), Liaison (B), iFlash (C), Euroimmun (D) and total antibody levels of Wantai (E) are reported after Log10 scale transformation. Patients/Subjects categories: blood donors (Pre-COV), negative autoimmune patients (AI), negative healthcare workers (NegHW), SARS-CoV-2 positive patients, subdivided into Mild (Mild), Moderate (Mod) and Severe (Sev) symptoms. Statistically significant differences: for Maglumi and iFlash, Pre-COV, AI and NegHW vs SARS-CoV-2 positive patients (p<0.01 for all); for Liaison, Pre-COV, AI and NegHW vs SARS-CoV-2 positive patients (p<0.01 for all) and Mild vs Sev (p = 0.017); for Euroimmun, AI and NegHW vs SARS-CoV-2 positive patients (p<0.01 for all) and Mild vs Sev (p = 0.006); for Wantai, p = not significant for all.

**Figure 2.**
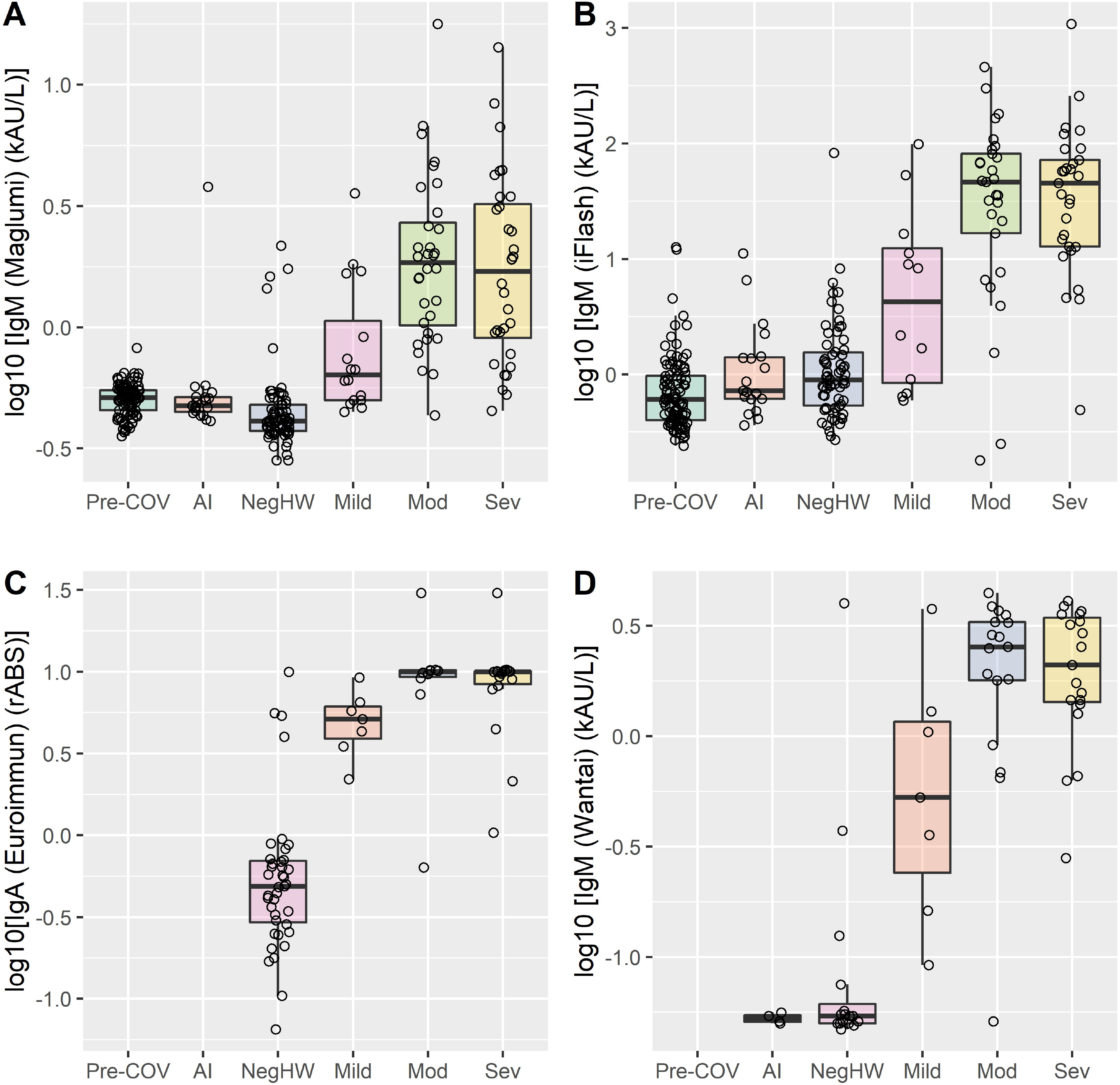
– IgM antibody levels are reported for Maglumi (A), iFlash (B) and Wantai (D). IgA antibody levels are reported for Euroimmun (C). Log10 scale transformation was used. Blood donors (Pre-COV), negative autoimmune patients (AI), negative healthcare workers (NegHW), SARS-CoV-2 positive patients, subdivided into Mild (Mild), Moderate (Mod), and Severe (Sev) symptoms. Statistically significant differences: for Maglumi, Pre-COV and AI vs NegHW and SARS-CoV-2 positive patients (p<0.01 for all) and Mild vs Mod and Sev (p<0.01 and p = 0.022, respectively); iFlash, Pre-COV and AI vs NegHW and SARS-CoV-2 positive patients (p<0.01 for all) and Mild vs Sev (p = 0.041, respectively); for Euroimmun, NegHW vs Sev (p = 0.009) and Mild vs Sev (p=0.044); for Wantai, NegHW vs SARS-CoV-2 positive patients (p=0.03 for Mild, p < 0.01 for Mod and Sev).

**Table 2.** - Maglumi, Liaison, iFlash diagnostic characteristics, considering positive or negative SARS-Cov-2 swab test in positive and negative patients (with onset > 11 gg).

| Analytical Systems | N | AUC<br>(95% CI)<br>% | Redefined thresholds |  |  |  |  |  | Manufacturer threshold |  |  |  |  |  |
| --- | --- | --- | --- | --- | --- | --- | --- | --- | --- | --- | --- | --- | --- | --- |
|  |  |  | Optimized<br>Cut-off* | Sensitivity<br>% | Specificity<br>% | LR+<br>(95% CI) | LR-<br>(95% CI) | Classification<br>accuracy | Cut-off | Sensitivity<br>% | Specificity<br>% | LR+<br>(95% CI) | LR-<br>(95% CI) | Classification<br>accuracy |
| Maglumi | 170 | 96.2<br>(93.4-99.0) | 2.0 | 93.8<br>(86.0-97.9) | 93.3<br>(86.1-97.5) | 29.84<br>(13.55-65.73) | 0.06<br>(0.03-0.15) | 93.53%,<br>C-k = 0.870<br>(SE=0.07) | 1.1 | 95.0<br>(87.7-98.6) | 88.9<br>(80.5-94.5) | 8.55<br>(4.76-15.37) | 0.06<br>(0.02-0.15) | 91.76%<br>C-k = 0.835<br>(SE =0.06) |
| Liaison | 131 | 96.6<br>(93.1-100) | 6.2 | 97.1<br>(89.8-99.6) | 88.9%<br>(78.4- 95.4) | 8.74<br>(4.34-17.58) | 0.03<br>(0.01-0.13) | 93.13%,<br>C-k = 0.862<br>(SE=0.09) | 15 | 82.4<br>(71.2-90.5) | 96.8<br>(89.0-99.6) | 25.94<br>(6.60-101.90) | 0.18<br>(0.11-0.31) | 89.31%<br>C-k=0.787,<br>SE=0.08 |
| iFlash | 156 | 95.9<br>(92.3-99.5) | 15.0 | 97.2%<br>(90.2-99.7) | 85.9<br>(876.6-92.5) | 6.88<br>(4.07-11.65) | 0.03<br>(0.01-0.13) | 91.0%,<br>C-k = 0.821<br>(SE=0.08) | 10 | 93.0<br>(84.3-97.7) | 92.9<br>(85.3-97.4) | 13.17<br>(6.07-28.56) | 0.08<br>(0.03-0.18) | 92.95%,<br>C-k=0.858<br>(SE=0.08) |

\*Youden index; C-k = Cohen's kappa

#### Performances of CLIA methods for IgG Ab

Different numbers of samples were measured for each assay, depending on the availability of reagents, and in particular, 170 for Maglumi, 131 Liaison and 156 for iFlash. The ROC analyses underlined overlapping results in terms of AUC for all assays.

Following the manufacturers’ specifications, the sensitivities, specificities, likelihood ratios (LR), classification accuracy and Cohen’s kappa were calculated and reported (Table 2). The highest sensitivity and specificity were obtained for Maglumi and Liaison, respectively. The performances of the two assays resulted in a negative and positive likelihood ratio of 0.06 and 25.94, respectively. Classification accuracies were greater than 90% for Maglumi and iFlash.

Using the Youden index metric, for each assay the best thresholds were estimated. These thresholds were different from manufacturers’ suggested cut-offs, especially for Maglumi and Liaison. The redefined thresholds allowed higher values to be obtained for: specificity, classification accuracy and positive LR for Maglumi; sensitivity, accuracy and negative LR for Liaison; sensitivity and negative LR for iFlash. Using these redefined thresholds, the predictive characteristics of each assay were investigated by Fagans’ nomogram considering the prevalence of disease detected among healthcare workers at the University-Hospital of Padova as 0.04 (4%; data not shown). The results showed that Liaison and iFlash assays allowed an almost perfect classification of negative subjects, with a post-test probability of not-having a disease of around 0.0015 (0.15%) (Supplementary Fig. 1).

#### Agreement of CLIA and ELISA assays

The pairwise agreements between the results of CLIA and ELISA assays were evaluated considering a total of 79 samples for the comparison of IgG obtained on Maglumi, Liaison, iFlash and Euroimmun. Sixty-three of the 79 samples were used for the comparison between Wantai AbT and the other assays, and for overall agreement. Supplementary Figure 2 shows the results obtained with Bland Altman analysis and Passing Bablok regressions for CLIA assays. Concordance was calculated on positive/negative assay results using the thresholds from the Youden index for CLIA, and from manufacturers for ELISA. The greatest agreements were obtained for Liaison/Euroimmun (Cohen’s kappa = 0.945), Liaison/Wantai AbT (Cohen’s kappa = 0.961), Euroimmun/Wantai AbT (Cohen’s kappa = 0.962), with a percentage of concordant results of more than 97 (Table 3). Overall agreement was 90.3% (Cohen’s kappa = 0.805 and SE = 0.041) for CLIA, 98.4% (Cohen’s kappa = 0.962 and SE = 0.126) for ELISA.

**Table 3.**
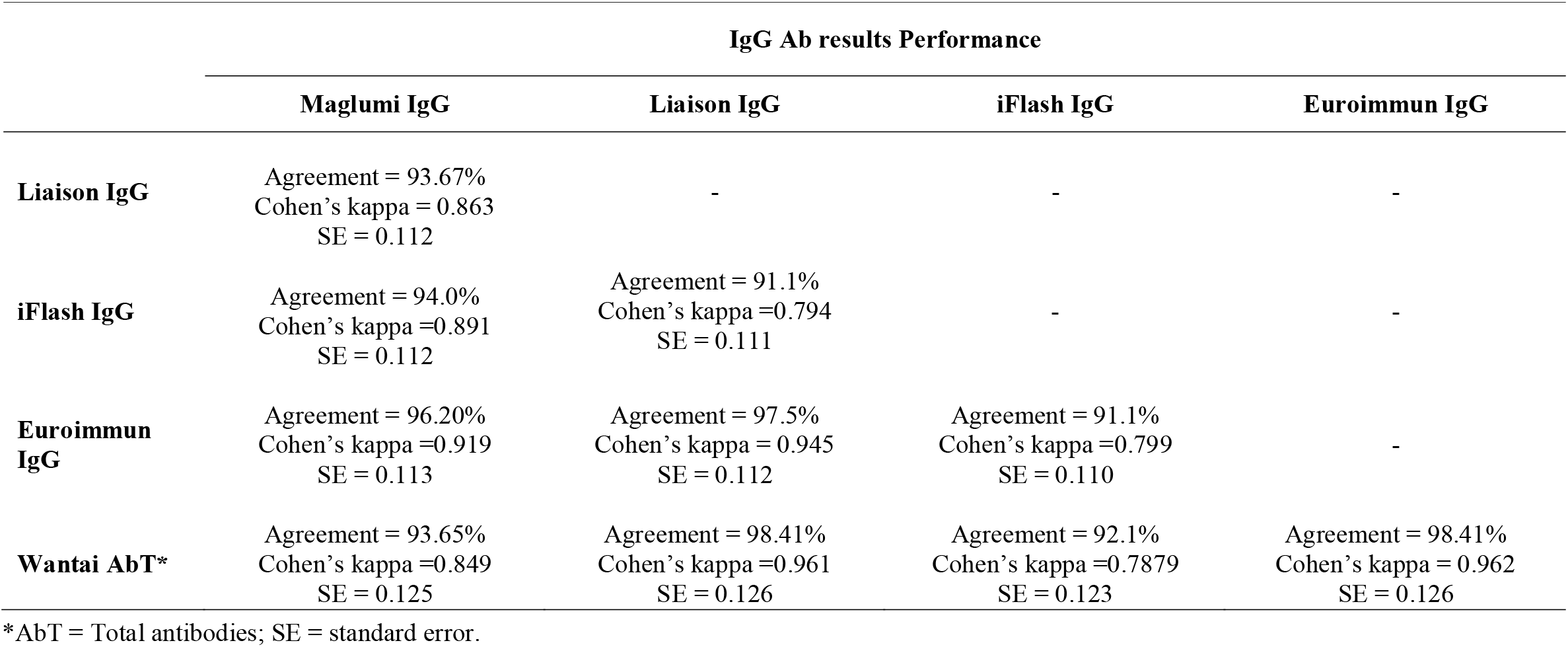
– Agreement and Cohen’s kappa of the 5 analytical systems under evaluation, using the best cut-off from Youden index for CLIA methods and manufactures’ defined cut-off for Euroimmun and Wantai AbT. Seventy-nine samples were used for the comparison, only 63 samples for the Wantai AbT assay.

| IgG Ab results Performance |  |  |  |  |
| --- | --- | --- | --- | --- |
|  | Maglumi IgG | Liaison IgG | iFlash IgG | Euroimmun IgG |
| **Liaison IgG** | Agreement = 93.67%<br>Cohen’s kappa = 0.863<br>SE = 0.112 | - | - | - |
| **iFlash IgG** | Agreement = 94.0%<br>Cohen’s kappa =0.891<br>SE = 0.112 | Agreement = 91.1%<br>Cohen’s kappa =0.794<br>SE = 0.111 | - | - |
| **Euroimmun IgG** | Agreement = 96.20%<br>Cohen’s kappa =0.919<br>SE = 0.113 | Agreement = 97.5%<br>Cohen’s kappa = 0.945<br>SE = 0.112 | Agreement = 91.1%<br>Cohen’s kappa =0.799<br>SE = 0.110 | - |
| **Wantai AbT\*** | Agreement = 93.65%<br>Cohen’s kappa =0.849<br>SE = 0.125 | Agreement = 98.41%<br>Cohen’s kappa =0.961<br>SE = 0.126 | Agreement = 92.1%<br>Cohen’s kappa =0.7879<br>SE = 0.123 | Agreement = 98.41%<br>Cohen’s kappa = 0.962<br>SE = 0.126 |

\*AbT = Total antibodies; SE = standard error.

#### Definition of thresholds harmonizing IgG Ab CLIA assays results

The analyses made enabled us to define the best (possible) threshold for allowing agreement of CLIA results. The highest agreements and Cohens’ kappa were achieved with the following assays/thresholds settings:

a. 92.9% and 0.856 for Maglumi and Liaison with 2.0 kAU/L and 7.6 kAU/L cut-offs, respectively;
b. 97.2% and 0.944 for Maglumi and iFlash with 1.6 kAU/L and 10 kAU/L cut-offs, respectively;
c. 94.2% and 0.883 for Liaison and iFlash with 7.6 kAU/L and 15.8 kAU/L cut-offs, respectively. Considering all CLIA assays, the highest agreements and Cohens’ kappa were achieved with the following threshold settings: 91.0% and 0.879 for Maglumi at 2.0 kAU/L, Liaison at 7.6 kAU/L and iFlash with 15.8 kAU/L.

### Discussion

There is an urgent need to identify strategies aiming to safely ease lockdown measures, thus allowing a return to productive economic levels, and social activity. Individuals who test positive for antibodies against SARS-CoV-2 could act as ‘shields’ against transmission. A recent model suggested that serological testing could make an important contribution in the reduction of viral spread and overall mortality (9). Rigorous comparative performance data are crucial to understanding the potential clinical usefulness of serological assays, starting from the evaluation of analytical performance characteristics to improve the definition of diagnostic accuracy not only in terms of specificity and sensitivity but also as positive and negative likelihood ratios, in order to provide reliable clinical information in different disease prevalence settings. A significant challenge in determining whether an individual is immune to SARS-CoV-2 depends on the fact that, so far, serologic data have mainly been obtained in hospitalized symptomatic patients. Serologic findings in asymptomatic or mildly symptomatic exposures may not present the same high degree of correlationin hospitalized patients. Therefore, we evaluated not only hospitalized patients but also healthcare operators in order to provide evidence of any different antibody behavior. Currently, there is an urgent need to identify those subjects who were not previously infected by COVID-19, in order to prevent further spread of the virus. In this study we focused on detecting the virus in healthcare workers, who are at a high risk of contracting the disease and consequently putting patients and coworkers at risk. For this purpose, we performed an in-depth optimization of assays thresholds, to achieve the best negative likelihood ratios. COVID-19 patients were identified according to the currently recognized “gold standard”: a positive nasopharyngeal swab test result for both hospitalized patients and healthcare workers. In view of current knowledge of seroconversion time and antibody kinetics (10,11), we only included samples collected after 11 days from the onset of symptoms. SARS-CoV-2 negative healthcare workers were defined as subjects with at least three sequential negative nasopharyngeal swab test as a criterion to assure the complete absence of infection. In addition, we included 101 donors with samples collected in 2015 (before the emergence of SARS-CoV-2) and 19 autoimmune patients in order to verify possible analytical interferences.

At the time the study started, the data available on precision for CLIA assays were limited. Apart from Maglumi, which we evaluated in a previous study, Liaison and iFlash assays repeatabilities and intermediate precisions were estimated and results obtained with both methods proved satisfactory. Figure 2 shows the comparison data for IgM and IgA SARS-CoV-2 antibodies. The Liaison and Euroimmun assays have been developed to measure SARS-CoV-2 IgG only, and therefore IgM were not available, whilst Euroimmun IgA was evaluated and included in the comparison. As shown, and in agreement with findings reported in recently published studies (12), IgM does not provide valuable information for study purposes, and therefore these results were not included in the assays comparison. Interestingly, as highlighted in Figure 2, some Neg-HW cases were found to be positive whereas some SARS-CoV-2 positive patients were negative in all IgG, and IgM and IgA, methods. This might suggest that some SARS-CoV-2 positive patients neither produce detectable IgG antibodies, nor produce IgA and IgM, even if the nasopharyngeal swab test is positive.

The overall performances of assays were elevated, AUC being above 96% for all the three CLIA methods, although overlapping confidence intervals show that this finding was not of statistical significance. Further studies are required to confirm this observation. The threshold redefinition was effective in improving diagnostic performances. Considering the purpose of achieving the best negative likelihood ratio, the optimized cut-offs allowed us to obtain a real improvement for Liaison and iFlash. Furthermore, the Fagan’s nomogram was used to enhance the provision of evidence of the usefulness of IgG values in clinical practice, emphasizing achievements. Given a pre-test probability of disease (e.g. 0.04 or 4%), this tool shows the post-test disease probability for both positive and negative test results. In an ongoing study of the Veneto Region, the estimated prevalence of COVID-19 disease is 4% (data not shown). Consequently, the final probability of a healthcare worker with a negative test result contracting COVID-19 infection is around 0.15% for Liaison and iFlash and 0.3% for Maglumi; this, from a clinical view-point means that a negative result provides a highly satisfactory exclusion power.

The between-methods agreements obtained were all above 93%, but our data suggest that the highest agreements can be achieved with ELISA assays. A combination of thresholds was estimated in order to improve the overall agreement between CLIA assays. The thresholds calculated allowed us to obtain the highest agreement (91%) on adopting the threshold that assured the best performance (Youdex index) for Maglumi and Liaison, while for iFlash the threshold was higher than that identified with the Youden index, as previously reported. This, in turn, provides preliminary evidence that analytical harmonization is not a “mission impossible”.

The present study has some limitations. First, antibody dynamics monitoring was extended to a maximum of 54 days, the initial COVID-19 patients being admitted to our hospital at the end of February 2020. Second, the limitation in sample sizes and reagents precluded measurement with different assays in the same number of subjects. Third, the relationships of currently measured antibodies with neutralizing activity against SARS-CoV-2 were not evaluated. A body of evidence, however, demonstrates that antibodies targeting different domains of S protein, including S1, RBD and S2, may all contribute to virus neutralization (13, 14).

### Conclusions

Overall performances of the evaluated CLIA assays were highly satisfactory, and allowed us to achieve an accurate classification. Moreover, good agreement was found between CLIA and ELISA assays. The results obtained indicate that it might be possible to define thresholds that improve the negative likelihood ratio. On considering healthcare workers, a negative test result allowed us to identify negative subjects, who should then be closely monitored over time to prevent viral spread.

## Data Availability

Data are available on requests, if the journal policy will allowed that.

## Acknowledgments

We thank Dr. Chiara Cosma and Daniela Rinaldi (medical laboratory Scientists) for their valuable technical support. We acknowledge DiaSorin, Euroimmun and Pantec (Yhlo distributor in Italy) for kindly supplying reagents without any influence in study design and data analysis.

## supplementary Figures

**Supplementary Figure 1:** Fagan’s nomograms for CLIA methods, calculated using the likelihood ratios obtained considering the thresholds from Youden’s index, and a prevalence of SARS-CoV-2 infection of 0.04 (4%). Post-test probabilities for positive (LR_positive) and negative (LR_Negative) test results are shown for A) Maglumi, B) Liaison and C) iFlash.

**Supplementary Figure 2:** Bland Altman and Passing Bablok analyses for: A) and D) Liaison vs Maglumi, B) and E) for Liaison and iFlash; C) and F) for iFlash and Maglumi.

## References

1. Cheng MP, Papenburg J, Desjardins M, Kanjilal S, Quach C, Libman M, et al. Diagnostic testing for Severe Acute Respiratory Syndrome-Related Coronavirus-2: A Narrative Review. Ann Intern Med 2020 (ahead of print) doi: 10.7326/M20-1301.

2. Mizumoto K, Kagaya K, Zarebsky A, Chowell G. Estimating the asymptomatic proportion of coronavirus disease 2019 (COVID-19) cases on board the Diamond Princess cruise ship, Yokohama, Japan, 2020. Euro Surveill 2020;25:10.

3. Binnicker MJ. Emergence of a Novel Coronavirus Disease (COVID-19) and the Importance of Diagnostic Testing: Why Partnership between Clinical Laboratories, Public Health Agencies, and Industry Is Essential to Control the Outbreak. Clin Chem 2020;66:664–6.

4. Sethuraman N, Jeremiah SS, Ryo A. Interpreting Diagnostic Tests for SARS-CoV-2. JAMA 2020, (ahead of print) doi: 10.1001/jama.2020.8259.

5. Long QX, Liu BZ, Deng HJ, Wu GC, Deng K, Chen YK, et al. Antibody responses to SARS-CoV-2 in patients with COVID-19. Nat Med 2020, (ahead of print) doi: 10.1038/s41591-020-0897-1.

6. Lassauniere R, Frische A, Harboe ZB, Nielsen ACY, Fomsgaard A, Krogfeltet KA, et al. Evaluation of nine commercial SARS-CoV-2 immunoassays. Preprint at: http://doi.org/10.1101/2020.04.09.20056325 (2020).

7. Padoan A, Cosma C, Sciacovelli L, Faggian D, Plebani M. Analytical performances of a chemiluminescence immunoassay for SARS-CoV-2 IgM/IgG and antibody kinetics. Clin Chem Lab Med 2020 (ahead of print) doi: 10.1515/cclm-2020-0443.

8. Padoan A, Sciacovelli L, Basso D, Negrini D, Zuin S, Cosma C, et al. IgA-Ab response to spike glycoprotein of SARS-CoV-2 in patients with COVID-19: A longitudinal study. Clin Chim Acta 2020; 507: 164–6.

9. Kraay ANM, Nelson KN, Zhao C, Weitz JS, Lopman BA. Modeling serological testing to inform relaxation of social distancing for COVID-19 control. Preprint at: https://doi.org/10.1101/2020.04.24.20078576 (2020).

10. Zhao J, Yuan Q, Wang H, Liu W, Liao X, Su Y, et al. Antibody responses to SARS-CoV-2 in patients of novel coronavirus disease 2019. Clin Infect Dis. 2020, (ahead of print) doi: 10.1093/cid/ciaa344.

11. Guo L, Ren L, Yang S, Xiao M, Chang Yang F, et al. Profiling Early Humoral Response to Diagnose Novel Coronavirus Disease (COVID-19). Clin Infect Dis. 2020 Mar 21. pii: ciaa310. doi: 10.1093/cid/ciaa310. [Epub ahead of print].

12. Landry ML. Immunoglobulin M for Acute Infection: True or False? Clin Vaccine Immunol. 2016; 23: 540–5.

13. To KK, Tsang OT, Leung WS, Tam AR, Wu TC, Lung DC, et al. Temporal profiles of viral load in posterior oropharyngeal saliva samples and serum antibody responses during infection by SARS-CoV-2: an observational cohort study. Lancet Infect Dis 2020;20:565–74.

14. Jiang S, Hillyer C, Du L. Neutralizing Antibodies against SARS-CoV-2 and other Human Coronaviruses. Trends in Immunology 2020; 41: 355–359.

